# Comprehensive evaluation of the use of intravitreal injection of anti-vascular endothelial growth factor drugs in patients with fundus lesions based on real-world data

**DOI:** 10.1101/2024.04.01.24305180

**Authors:** Cheng-qun Chen, Li-ping Du, Qing Liu, Qing-hua Ren, Zhen-feng Zhu, Gui-fang Sun, Yu-shen Li, Yang Yang, Shu-zhang Du, Yue-dong Qi

## Abstract

The prevalence of fundus lesion-related diseases is increasing, which ophthalmic anti-VEGF drugs have become the drugs of choice for the treatment of fundus lesions diseases. To evaluate the clinical value of three ophthalmic anti-VEGF drugs in the treatment of fundus lesions diseases, to guide the rational use of the clinic. Inpatients with fundus lesions who had intravitreal injections of Aflibercept, Conbercept and Leizumab during 2020 were studied and six indicators were selected for a comprehensive evaluation. In terms of safety, Aflibercept, Conbercept, and Leizumab experienced adverse effects of elevated Intraocular Pressure (IOP). In terms of effectiveness, Leizumab was strong, that of Aflibercept was stronger and that of Conbercept was weaker. In terms of economic, there was no significant difference in the cost of Aflibercept, Conbercept and Leizumab and a significant difference in the total treatment cost and the cost of surgery. In terms of appropriateness, Aflibercept was more suitable than Conbercept, and there was no significant difference between Leizumab and Aflibercept. In terms of accessibility, Aflibercept, Conbercept and Leizumab were all accessible to urban residents in Henan Province. For rural people, these are unreachable. In terms of innovation, Aflibercep was the most innovative, followed by Leizumab and finally Conbercept. In terms of effectiveness and accessibility, Leizumab performed best compared to Aflibercept and Conbercept. In terms of accessibility and innovation, Aflibercept performed best compared to Conbercept and Leizumab. In terms of safety and economic, Aflibercept, Conbercept and Leizumab performed comparably.

## Introduction

The number of patients with age-related fundus diseases is increasing in China, becoming a major cause of irreversible visual impairment in the middle-aged and elderly population and seriously affecting the lives of patients(1). In the future, this burden will be exacerbated by the ageing of the population(2). With the introduction of anti-angiogenic therapies, significant progress has been made in the treatment of exudative or so-called wet age-related macular degeneration(3). Anti-VEGF drugs are increasingly being used in the clinical treatment of ophthalmologically related diseases, and multiple applications are becoming more common (4). They have also shown clear advantages in clinical application, but the changing dynamics of the medical and economic issues associated with this suggest that significant challenges lie ahead(5). A comprehensive evaluation of the economics as well as efficacy-related aspects of these drugs is therefore becoming increasingly important (6). Different approaches to drug therapy also carry a corresponding economic burden for patients, and changes in drug therapy can also lead to changes in the economic burden(7). Drugs in this category are currently marketed as Aflibercept(7), Conbercept(8) and Leizumab(9). This study evaluates the combined clinical value of three ophthalmic anti-VEGF agents generated in the treatment of diseases associated with fundus lesions based on real-world data. The results of this study will help patients and physicians, as well as pharmacists, to promote the clinical use of these drugs and benefit more patients based on the corresponding findings.

## Materials and methods

### Data collection

The study data were obtained from the electronic medical records of the hospital information system starting from 2020, and patients discharged from the First Affiliated Hospital of Zhengzhou University in Zhengzhou City, Henan Province, from January 2020 to December 2020 were collected, and the collected patients were followed up from 2021 to 2022. Collection criteria: patients who had received intravitreal injection treatment at the First Affiliated Hospital of Zhengzhou University during 2020. Inclusion criteria: (1) age ≥ 18 years; (2) clinical diagnosis of fundus-related disease; (3) all used any 1 or more drugs of Aflibercept, Conbercept, and Leizumab for the treatment of ophthalmic disease during hospitalization. Exclusion criteria: (1) patients explicitly did not agree to follow-up at the return visit; (2) data that were not part of a real-world study.

The total number of cases of Aflibercept, Conbercept, and Leizumab used by all inpatients in the study time cross-section was obtained, and the total number of cases was determined according to the inclusion and exclusion principles. Information was collected from the hospital medical record system, including hospitalisation number, age, gender, number of hospitalisations, primary diagnosis, other diagnoses, date of admission, date of discharge, economics data, data related to adverse drug reactions, and information on relevant drugs used to combat adverse reactions.

### Selection of main evaluation indicators

The study was based on the “Guidelines for the Management of Comprehensive Clinical Evaluation of Pharmaceuticals (Trial Version 2021)”, and the evaluation indicators were determined according to the characteristics of pharmaceutical applications in the real world. Aflibercept, Conbercept and Leizumab were comprehensively evaluated by six indicators including safety, efficacy, economy, suitability, accessibility and innovation. The accessibility of Aflibercept, Conbercept and Leizumab was evaluated specifically using the WHO/HAI standard survey method(10).

### Safety indicators

Safety indicators are included in the evaluation criteria, the amount of information related to adverse reaction reporting and adverse reaction events is low, in the process of adverse drug reaction reporting, the reporting rate of new serious adverse drug reactions is high, while the reporting rate of common and minor adverse reactions is low, the data collected through the adverse reaction reporting platform is very little, in order to improve the evaluation of drug safety, information on patient safety was analyzed and the incidence of common adverse reactions in practice with the three ophthalmic anti-VEGF drugs was analyzed on the basis of whether the use of Aflibercept, Conbercept and Leizumab was followed by the use of drugs to combat the symptoms of related adverse reactions. Adverse effects of elevated IOP as a test for common adverse effects because this adverse effect is not susceptible to the patient’s own illness and disease progression. If other indicators need to be evaluated the data can be mined in depth.

### Effectiveness indicators

By analyzing the number of days between two ophthalmic anti-VEGF drug injections in patients, the longer interval is considered to be a case where the drug has a strong therapeutic effect on the disease and thus the effectiveness of that ophthalmic anti-VEGF drug. The number of drug changes and the number of cases changed were counted, and the data were finally enumerated based on the discharge dates of patients in the real world data to obtain the hospitalization intervals, as the reasons for drug changes may be influenced by many aspects such as the physician’s personal medication habits, drug availability and the actual situation during treatment. To ensure a more accurate evaluation of the effectiveness of each of the three ophthalmic anti-VEGF drugs, patients who had been hospitalised more than twice but had changed ophthalmic anti-VEGF drugs were excluded from the effectiveness evaluation, and only patients who had been hospitalised twice or more and had not changed ophthalmic anti-VEGF drugs were included in the evaluation of effectiveness indicators.

### Economic indicators

The economy of Aflibercept, Conbercept and Leizumab was evaluated by statistically analyzing the cost of the patient’s hospitalisation and the total hospitalisation cost to assess whether there was a significant difference between the three ophthalmic anti-VEGF drugs. Patients’ surgical costs were analyzed against the costs of Aflibercept, Conbercept and Leizumab to evaluate whether there was a significant difference.

### Appropriateness indicators

The appropriateness of these three ophthalmic anti-vascular drugs was assessed based on indications in real-world data for Aflibercept, Conbercept and Leizumab and information on drug indications in the drug formulary. When doctors use the anti-VEGF drugs clinically, they diagnose the patient, default the diagnosis stated by the doctor to be correct, analyze whether the indications for Aflibercept, Conbercept and Leizumab comply with the drug indication when used in the real world based on the diagnosis, and the number of cases meeting the drug indications was also compared with the number of cases using the three drugs mentioned above and a chi-square analysis was used to derive the difference in suitability between Aflibercept, Conbercept and Leizumab.

### Accessibility indicators

The accessibility of the three ophthalmic anti-VEGF drugs was evaluated based on real world data characteristics. The evaluation was carried out using the per capita income assessment burden of disease method. The accessibility of the three ophthalmic anti-VEGF drugs was evaluated by analyzing the total cost of the drug spent on a standard dose of the drug to treat a disease over a course of treatment, which was equivalent to a multiple of daily disposable income per capita. The multiples of Aflibercept, Conbercept and Leizumab equivalent to urban and rural per capita disposable income were calculated based on the data of urban and rural disposable income in the Henan Provincial Statistical Yearbook 2021 published by the Statistical Bureau of Henan Province, China and the cost per course of treatment of Aflibercept, Conbercept and Leizumab in real world data.

### Indicators of innovativeness

Review the relevant literature to obtain the time on the market and the respective patent status of Aflibercept, Conbercept and Leizumab to evaluate the innovativeness of the above three ophthalmic anti neovascular drugs. The evaluation was based on the number of patents.

### Statistics

Different statistical analysis methods were adopted according to the characteristics of the data, including one-way ANOVA and multivariate descriptive statistics.

## Results

### Patient characteristics

According to the inclusion criteria of this study, a total of 1292 patients at the First Affiliated Hospital of Zhengzhou University had used at least one of the three drugs, Aflibercept, Conbercept and Leizumab, between January 2020 and December 2020, of which 200 patients used Aflibercept, 311 patients used Conbercept, 665 patients used Leizumab, 112 patients used one of 1 and then switched to the other, and 4 patients used all 3 ophthalmic anti-VEGF drugs.

### Safety

There were a certain number of cases in the real-world data in which Aflibercept, Conbercept and Leizumab were followed by three IOP-lowering drugs, brimonidine, brinzolamide/timolol and brinzolamide, to control the symptoms of elevated IOP following intravitreal injection of ophthalmic anti-VEGF drugs. The number of cases in which the IOP-lowering drugs brimonidine, brinzolamide \ timolol and brinzolamide were used after injections of Aflibercept, Conbercept and Leizumab was: 49 out of 200 cases in which the IOP-lowering drugs were used in patients who had been given Aflibercept (Fig 1). Of the 311 patients who had used Conbercept, a total of 81 patients used of IOP-lowering medication (Fig 2). Of the 665 patients who used Leizumab, a total of 148 patients used an IOP-lowering drug (Fig 3).

**Figure 1.**
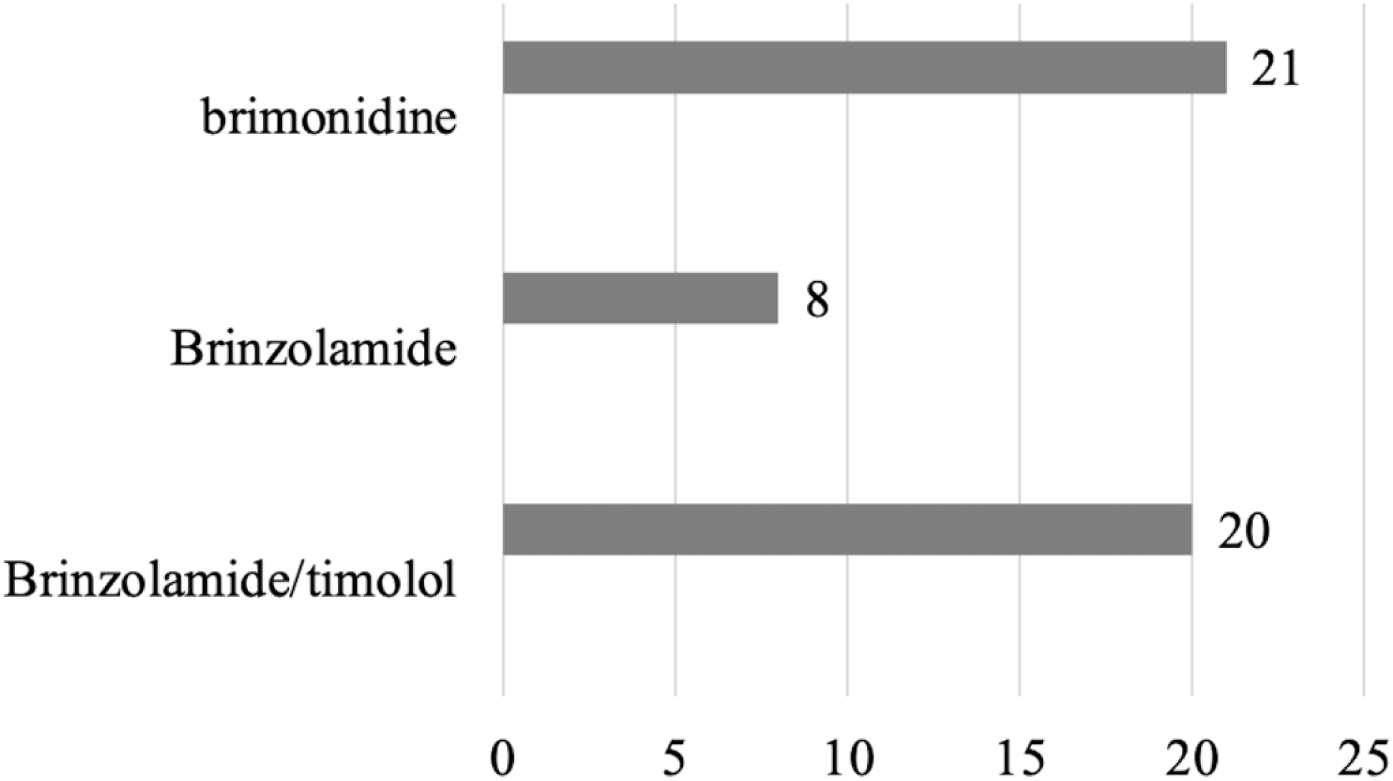
The use of three IOP-lowering drugs after the use of Aflibercept.

**Figure 2.**
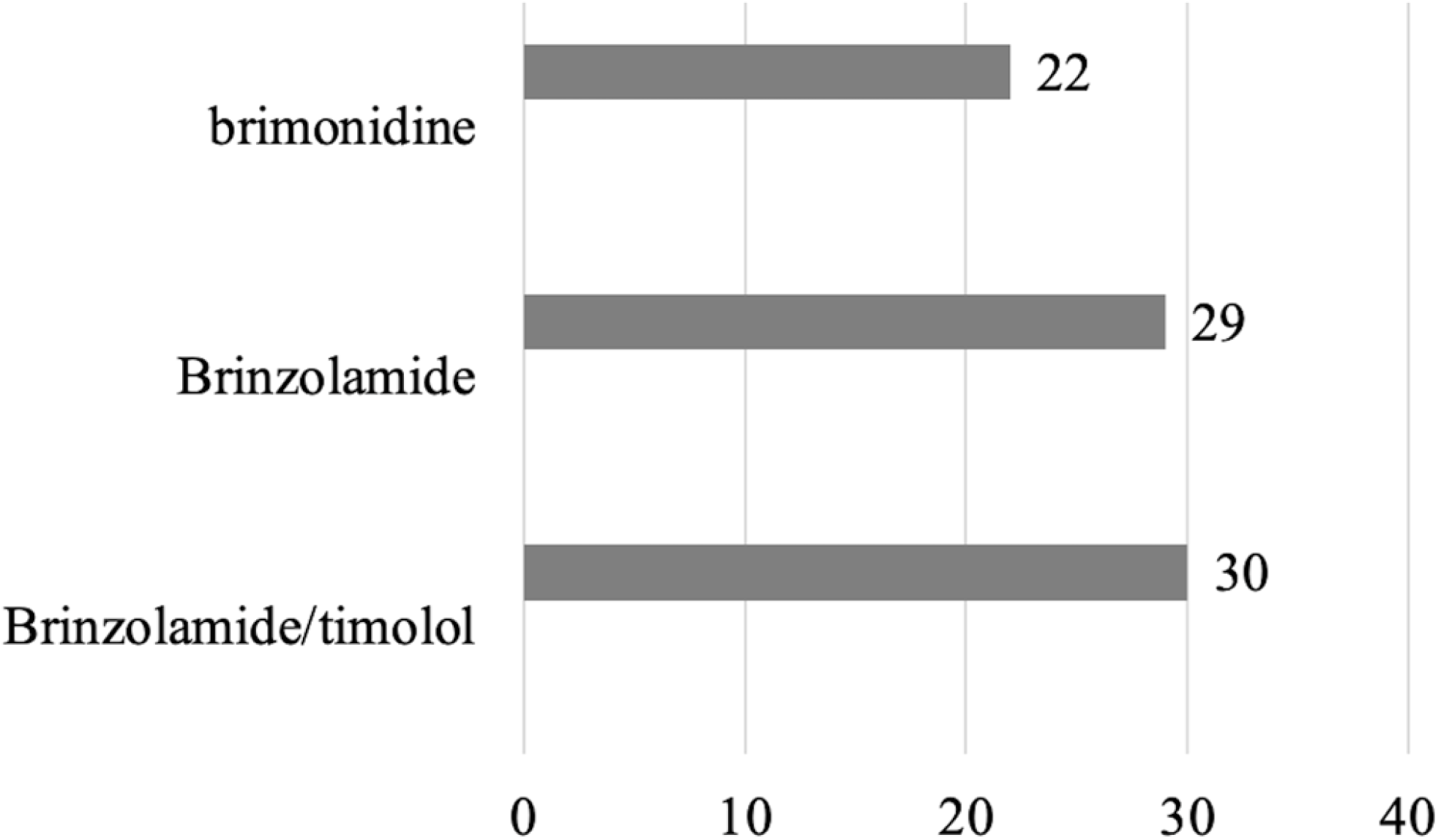
The use of three IOP-lowering drugs after the use of Conbercept.

**Figure 3.**
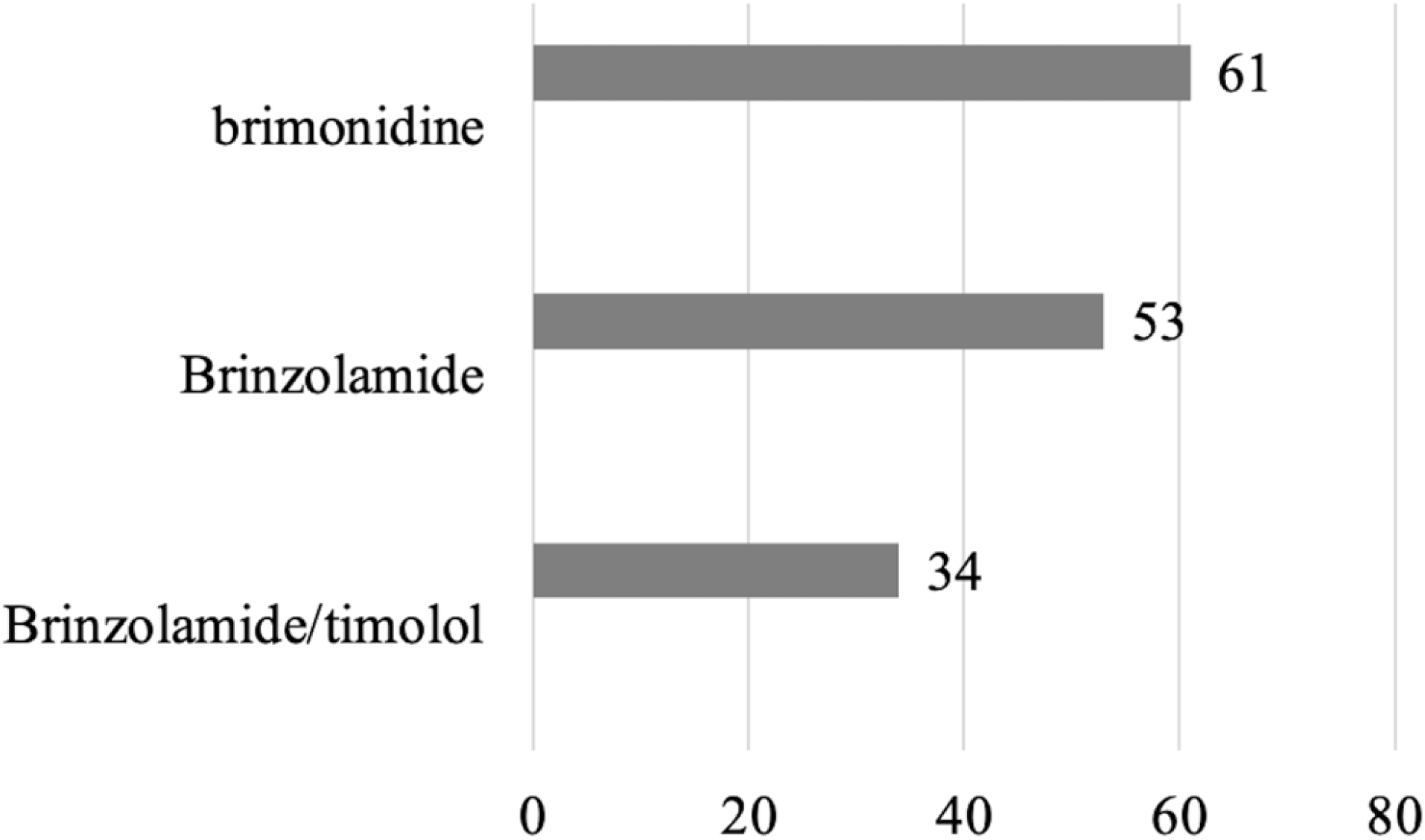
The use of three IOP-lowering drugs after the use of Leizumab.

### Effectiveness

Cases that used only one of Aflibercept, Conbercept and Leizumab and were hospitalised twice or more were analyzed according to the basic characteristics of all cases included in the study and the mean number of days in hospital for such cases was calculated. Calculations showed that the mean number of days between hospitalisations was 56.6 days for patients on Aflibercept, 49.3 days for those on Conbercept and 59.6 days for those on Leizumab.

### Economics

After analyzing the cost and total cost of the three ophthalmic anti-VEGF drugs and the cost of surgery in patients using anti-VEGF drugs for ocular injections, the statistical results were that there was no statistical difference between Aflibercept and Conbercept (p>0.05), Aflibercept and Leizumab (p>0.05) and There was no statistical difference in cost between Conbercept and Leizumab (p>0.05) Table 1.

**Table 1.** Comparison of the cost of three anti-VEGF drugs with the total cost of hospitalisation.

| Dependent Variable | (I)Drug Codes | (J)Drug Codes | Mean Different(I-J) | Std.Error | Sig. |
| --- | --- | --- | --- | --- | --- |
| Western medicine fees | Aflibercept | Conbercept | -476.65 | 282.01223 | 0.091 |
|  |  | Leizumab | -267.91 | 250.792 | 0.286 |
|  | Conbercept | Aflibercept | 476.65 | 282.01223 | 0.091 |
|  |  | Leizumab | 208.74 | 213.35447 | 0.328 |
|  | Leizumab | Aflibercept | 267.91 | 250.792 | 0.286 |
|  |  | Conbercept | -208.74 | 213.35447 | 0.328 |
| Total cost | Aflibercept | Conbercept | -3576.96* | 882.97479 | 0 |
|  |  | Leizumab | -1738.99* | 776.68559 | 0.025 |
|  | Conbercept | Aflibercept | 3576.96* | 882.97479 | 0 |
|  |  | Leizumab | 1837.97* | 725.61256 | 0.011 |
|  | Leizumab | Aflibercept | 1738.99* | 776.68559 | 0.025 |
|  |  | Conbercept | -1837.97* | 725.61256 | 0.011 |

\*. The level of significance of the difference in means is 0.05.

In terms of the total cost of the three ophthalmic anti-VEGF drugs, there was a statistical difference between the total cost of Aflibercept and Conbercept (p<0.05), the total cost of Aflibercept and Leizumab (p<0.05) and the total cost of Conbercept and Leizumab (p<0.05). The statistical results between the surgical costs of the three ophthalmic anti-VEGF drugs were statistically different between the surgical costs of Aflibercept and Conbercept (p < 0.05) Table 2. There was no statistical difference between the cost of surgery for Aflibercept and Leizumab (p > 0.05). There was a statistically significant difference between the cost of surgery for Conbercept and Leizumab (p < 0.05). The significance level of the difference in means was 0.05.

**Table 2.** Comparison of the cost of three anti-VEGF drugs with the cost of surgery.

| Dependent Variable |  |  | Mean Difference (I-J) | Std. Error | Sig. |
| --- | --- | --- | --- | --- | --- |
| Operation Fee | Aflibercept | Conbercept | -922.35* | 229.58148 | 0.000 |
|  |  | Leizumab | -69.22 | 204.16561 | 0.735 |
|  | Conbercept | Aflibercept | 922.35* | 229.58148 | 0.000 |
|  |  | Leizumab | 853.12* | 173.68834 | 0.000 |
|  | Leizumab | Aflibercept | 69.22 | 204.16561 | 0.735 |
|  |  | Conbercept | -853.12* | 173.68834 | 0.000 |
| Medicine costs | Aflibercept | Conbercept | -476.65 | 282.01223 | 0.091 |
|  |  | Leizumab | -267.91 | 250.79200 | 0.286 |
|  | Conbercept | Aflibercept | 476.65 | 282.01223 | 0.091 |
|  |  | Leizumab | 208.74 | 213.35447 | 0.328 |
|  | Leizumab | Aflibercept | 267.91 | 250.79200 | 0.286 |
|  |  | Conbercept | -208.74 | 213.35447 | 0.328 |

\*. The mean difference is significant at the 0.05 level.

### Appropriateness

Classify the patient’s disease according to the primary diagnosis in the real world versus other diagnoses, code and record the classified diagnoses. The indications for Aflibercept, Conbercept, and Leizumab were analyzed for real-world application in accordance with the drug specification. Aflibercept intraocular injection is indicated for the treatment of neovascular (wet) age-related macular degeneration (nAMD) in adults; diabetic macular oedema (DME). Conbercept is indicated for the treatment of: neovascular (wet) age-related macular degeneration (nAMD); visual impairment due to choroidal neovascularisation (pmCNV) secondary to pathological myopia; visual impairment secondary to diabetic macular oedema (DME). Leizumab is used for the treatment of wet (neovascular) age-related macular degeneration (AMD), visual impairment due to diabetic macular oedema (DME), visual impairment due to macular oedema secondary to retinal vein occlusion (RVO) (branch retinal vein occlusion (BRVO) or central retinal vein occlusion (CRVO)), choroidal neovascularisation (CNV, i.e secondary to pathological myopia (PM) and other causes of CNV). After comparing the real-world indications for the three ophthalmic anti-VEGF drugs with those in the medication aid, statistical analysis showed that of the 200 patients who used Aflibercept, 138 had a primary diagnosis that matched the drug’s instructions and 62 had a primary diagnosis that differed from the drug’s instructions Table 3.

**Table 3.** Comparison of compliance with the indications for Aflibercept and Conbercept.

| Title | | Aflibercept | Conbercept | $\chi^2$ | p |
| --- | --- | --- | --- | --- | --- |
| Is it qualified | yes | 138 | 171 | 10.004 | 0.002 |
|  | no | 62 | 140 |  |  |
| Total |  | 200 | 311 |  |  |

A. 0 cells (0.0%) are expected to have counts less than 5. The minimum expected count is 79.06.

Of the 311 patients who used Conbercept, 171 patients had a primary diagnosis that was in accordance with the drug instructions and 140 patients had a primary diagnosis that was different from the drug instructions Table 4.

**Table 4.** Comparison of compliance with the indications for Aflibercept and Leizumab.

| Title | | Aflibercept | Leizumab | $\chi^2$ | p |
| --- | --- | --- | --- | --- | --- |
| Is it qualified | yes | 138 | 409 | 3.717 | 0.054 |
|  | no | 62 | 256 |  |  |
| Total |  | 200 | 665 |  |  |

A. 0 cells (0.0%) are expected to have counts less than 5. The minimum expected count is 73.53.

Of the 665 patients who used Leizumab, 409 patients had a primary diagnosis that was consistent with the drug insert and 256 patients had a primary diagnosis that was different from the drug insert Table 5.

**Table 5.** Comparison of compliance with the indications for Conbercept and Leizumab.

| Title | | Conbercept | Leizumab | $\chi^2$ | p |
| --- | --- | --- | --- | --- | --- |
| Is it qualified | yes | 171 | 409 | 3.736 | 0.053 |
|  | no | 140 | 256 |  |  |
| Total |  | 311 | 665 |  |  |

A. 0 cells (0.0%) are expected to have counts less than 5. The minimum expected count is 126.18.

There was a statistically significant difference (p<0.05) between Aflibercept and Conbercept in complying with the indications. There was no statistical difference (p>0.05) between Aflibercept and Leizumab in terms of indication compliance, and no statistical difference (p>0.05) between Conbercept and Leizumab in terms of indication compliance.

### Accessibility

Calculate the cost per course of treatment for Abciximab, Conbercept and Leizumab as a multiple of the per capita disposable income of urban and rural residents. For urban residents, the multiplier of per capita urban disposable income per course of treatment is 0.229 for Abciximab, 0.226 for Conbercept and 0.218 for Leizumab. For rural residents, a multiple of 1.068 per course of Abciximab, 1.053 per course of Conbercept and 1.014 per course of Leizumab is equivalent to the rural per capita disposable income. The World Health Organization considers a course of medication less than 1 times daily income as good affordability(11) Table 6.

**Table 6.**
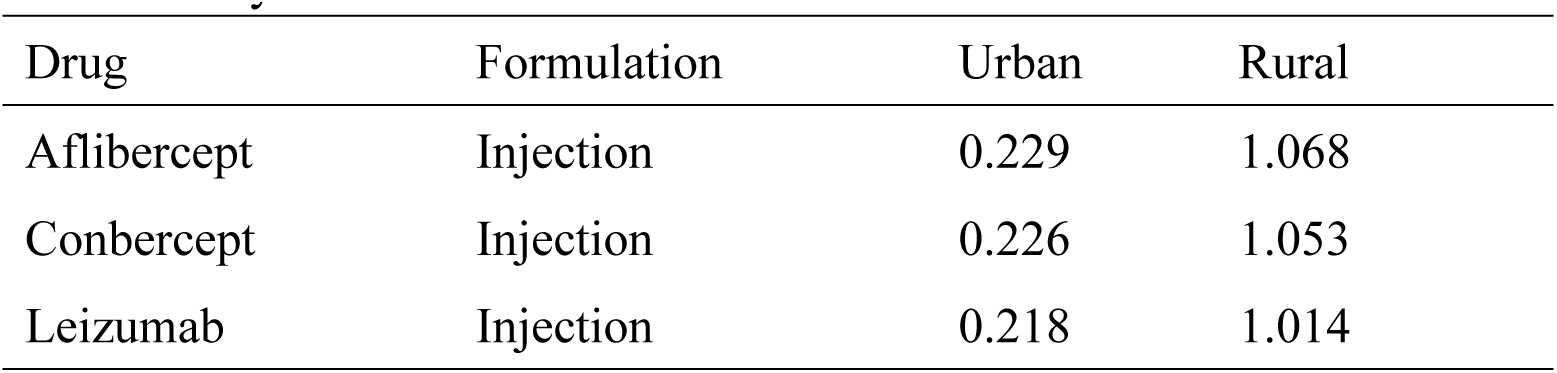
Urban/rural per capita daily disposable income evaluation affordability.

### Innovativeness

Aflibercept was approved for marketing by the FDA in 2011, and was launched in China in 2018. Conbercept was launched in China in 2013. Leizumab was approved for marketing by the FDA in 2006 and launched in China in 2018. The number of patents for Aflibercept is 33, 40, 33 for 2018-2020. the number of patents for Conbercept is 2, 3, 0. the number of patents for Leizumab is 24, 7, 25 respectively Table 7.

**Table 7.** Number of patents for three ophthalmic anti-VEGF drugs 2018-2020.

|  | Aflibercept | Conbercept | Leizumab |
| --- | --- | --- | --- |
| 2018 | 33 | 0 | 25 |
| 2019 | 40 | 3 | 7 |
| 2020 | 33 | 2 | 24 |
| Total | 106 | 5 | 56 |

## Discussion and conclusion

In this study, a multidimensional, systematic and scientific comprehensive evaluation of three ophthalmic anti-VEGF drugs was carried out, evaluating the three drugs in terms of safety, efficacy, economy, innovation, appropriateness and accessibility, with reference to the comprehensive clinical evaluation of the drugs. The study was conducted around the characteristics of comprehensiveness, with reliable sources of research data and more credible results. The results of the comprehensive evaluation of medicines conducted in this study can provide a reference for the clinical use process. The comprehensive evaluation of drugs emphasizes the comprehensive nature of the evaluation, including the evaluation content, evaluation methods and evaluation results.

The comprehensive evaluation of this study showed that, in terms of safety evaluation, all three ophthalmic anti-VEGF drugs experienced adverse effects of elevated IOP, which improved with the treatment of three different IOP-lowering drugs. The cause of IOP elevation during intravitreal injection of anti-VEGF drugs does not exclude the situation due to surgical manipulation, therefore it is important to examine the affected eye before administering IOP-lowering drugs and before proceeding with the next symptomatic treatment.

In terms of efficacy evaluation, the short dosing interval of Conbercept is relatively weak according to the efficacy evaluation criteria. Aflibercept has a longer dosing interval and is more effective. Leizumab has a long dosing interval and is highly effective. Based on the number of cases of the three ocular anti-VEGF drugs used in our hospital in 2020, the number of cases using Leizumab was the highest, which may be related to physicians’ dosing habits and the early availability of Leizumab.

In terms of economic evaluation, there was no significant difference in the cost of Aflibercept, Conbercept and Leizumab, and there was a significant difference in the total cost of all three. Analysis of the cost of surgery for all three revealed a significant difference between Aflibercept and Conbercept and a significant difference between Conbercept and Leizumab. The difference in surgical costs suggests that the frequency of neovascular drug treatment varies between patients depending on the diagnosis of the disease and the degree of disease progression. As there is no price level difference between Aflibercept, Conbercept and Leizumab in the overall treatment process, the other dimensions of the drug can be prioritized when choosing anti-VEGF drugs for the treatment of ophthalmic disease.

In terms of appropriateness assessment, Aflibercept is more suitable compared to Conbercept, there is no significant difference between Aflibercept and Leizumab, and there is no significant difference between Conbercept and Leizumab. When there are many off-label use cases of a drug in the real world, the research on the indication of the drug needs to be further strengthened. At the same time, clarification of the cause of the disease and the correct choice of ophthalmic anti-VEGF drugs during treatment will ensure optimal efficacy of the drug. It is important to avoid the irrational use and abuse of ophthalmic anti-VEGF drugs.

In terms of accessibility evaluation, the affordability of Aflibercept, Conbercept and Leizumab appears significantly different for urban and rural residents. For the urban population, Aflibercept, Conbercept and Leizumab were all affordable. The affordability of Leizumab is the strongest. Aflibercept is relatively the least affordable. Aflibercept, Conbercept and Leizumab cannot be afforded to rural residents. Ranibizumab was relatively less burdensome, and aflibercept was the most difficult to bear. According to the per capita disposable income of urban residents, three ophthalmic anti-VEGF drugs can be affordable, while the cost of three ophthalmic anti-VEGF drugs for rural residents will be unaffordable.

In terms of innovation evaluation, the patent value of Leizumab is medium, inferior to that of Aflibercept and superior to that of Conbercept.

A thorough understanding of the characteristics of the three drugs will help in the selection of clinical use.

## Data Availability

The data are owned by a third party and authors do not have permission to share the data.

## Acknowledgements

We thank Yu-shen Li and Yang Yang for their informational support. No potential conflict of interest was reported by the author(s).

